# Cumulative remnant cholesterol and major adverse cardiovascular events among adults with type 2 diabetes

**DOI:** 10.1101/2025.02.16.25322349

**Authors:** Yu-Wen Qian, Zhong-Yue Liu, Fei Fang, Jia-Min Wang, Xiao-Ping Shao, Yi-Ping Jia, Xian-Bo Wu, Fu-Rong Li, Wei Zhang, Huan-Huan Yang, Guo-Chong Chen, Hai-Peng Wang

**Author notes:** Corresponding to: Guo-Chong Chen, PhD, Department of Nutrition and Food Hygiene, School of Public Health, Suzhou Medical College of Soochow University, 199 Ren’ai Road, Suzhou 215123, China. OR Hai-Peng Wang, MD, Department of Cardiology, The First Affiliated Hospital of Soochow University, 188 Shizi Street, Suzhou 215006, China.

## Abstract

**Background:** Individuals with type 2 diabetes are prone to dyslipidemia. The relationship of cumulative exposure to elevated remnant cholesterol (RC) with major adverse cardiovascular events (MACEs) remains unclear for individuals with type 2 diabetes, especially for those with intensive lipid-lowering treatment.

**Methods:** This study included 5143 participants with type 2 diabetes from the Action to Control Cardiovascular Risk in Diabetes trial, who had at least four lipid measurements over the first three years of the trial and did not have MACEs across the measurements. Cumulative RC (cumRC) was calculated considering RC levels at each measurement and the between-measurement time interval. Cox proportional hazards regression models were used to estimate HRs and 95% CIs of MACEs associated with cumRC levels.

**Results:** During a median follow-up of 6.3 years, 472 participants developed MACEs, including 154 cardiovascular deaths, 211 with nonfatal myocardial infarction, and 148 with nonfatal stroke. After multivariable adjustment for conventional risk factors, higher levels of cumRC were associated with a higher risk of MACEs (HR_Q4 vs. Q1_ = 1.71, 95% CI: 1.31-2.22; P-trend <0.001), with similar associations for cardiovascular death (P-trend = 0.009) and nonfatal myocardial infarction (P-trend <0.001), but no association for nonfatal stroke (P-trend = 0.099). The association of cumRC with the risk of MACEs was consistent across age, sex, and lipid trial assignment groups and remained after further adjusting for baseline RC.

**Conclusion:** Among adults with type 2 diabetes, cumulative exposure to elevated RC is associated with a higher risk of MACEs, even in the context of intensive lipid management.

## Introduction

The majority of adult-onset diabetes are type 2 diabetes, which is a complex disease not only commonly featured by insulin resistance, but also accompanied by a variety of metabolic abnormalities, including dysregulation of blood lipids. Lipid abnormalities, especially elevated levels of low-density lipoprotein cholesterol (LDL-C), are established risk factors for atherosclerotic cardiovascular disease (CVD) [1]. Randomized controlled trials have revealed strong trends toward a reduction in major cardiovascular events with decreasing LDL-C levels via statins, both in non-diabetes and diabetes populations [2–4]. For example, the Cholesterol Treatment Trialists Collaborators demonstrated a 21% reduction in major vascular events for each 1 mmol/L decrease of LDL-C levels by statin therapy among individuals with diabetes, which was similar to the effect observed for those without diabetes [3].

Despite the cardiovascular benefits from LDL-C lowering, recent studies have shown a relatively high incidence of recurrent cardiovascular events for patients whose LDL-C levels had been pharmacologically lowered to the targeted range [5]. Such residual cardiovascular risk may be potentially attributable to elevated levels of remnant cholesterol (RC) [6, 7]. RC is used to describe the cholesterol content of all triglyceride-rich lipoproteins, or cholesterol other than LDL-C or HDL-C (high-density lipoprotein cholesterol), and is composed of very-low-density lipoprotein cholesterol, intermediate density lipoproteins, and chylomicron remnants [8, 9]. Mounting evidence has suggested that elevated levels of RC may partially explain the risk of cardiovascular events and death that remains after targeted LDL-C concentrations are reached [10]. There is also evidence that RC may be more useful than LDL-C for predicting atherosclerosis and assessing cardiovascular risk [11].

The majority of the previous studies on the association between RC and cardiovascular outcomes have used a single measurement of RC without considering longitudinal variations. RC levels may be influenced by various factors such as aging, lifestyle modification, and pharmacological interventions, such that an isolated measurement is likely insufficient to capture the long-lasting cardiovascular effect [12]. This may particularly be the case for diabetes population who are prone to dyslipidemia and the related therapies. Visit-to-visit variability in RC levels has been associated with the risk of developing major adverse cardiovascular events (MACEs) among individuals with type 2 diabetes [13]. These observations underscore the necessity of considering multiple RC measurements over time to better characterize the cardiovascular impact of long-term exposure to elevated levels of RC. Therefore, using data from the Action to Control Cardiovascular Risk in Diabetes (ACCORD) trial, we conducted a post hoc analysis to assess the relationship between cumulative RC (cumRC) levels and the risk of MACEs among adults with type 2 diabetes.

## Methods

### Study design and participants

Participants in the ACCORD trial (ClinicalTrials.gov number, NCT00000620) were recruited between June 2001 and October 2005 at 77 sites across the U.S. and Canada. The trial enrolled 10,251 people with a mean age of 62 years who had type 2 diabetes for a median duration of 10 years, had a mean glycated hemoglobin (HbA1c) level of 8.3%, and had a history of CVD or its risk factors (i.e., hyperglycemia, dyslipidemia, or high blood pressure [BP]). Detailed descriptions of the study design, methods, and main results have been published [14]. Briefly, the participants were randomly assigned to receive intensive therapy targeting an HbA1c level below 6.0 % or standard therapy targeting a level of 7.0 to 7.9 %. Participants were also assigned to one of two BP interventions (intensive systolic BP target < 120 mmHg, or standard treatment target < 140 mmHg), or a lipid intervention (placebo or fenofibrate). All surviving ACCORD participants who could be contacted were subsequently invited to participate in the ACCORDION study, during which data on cardiovascular and other health-related outcomes and measurements were collected between May 2011 and October 2014. The ACCORD/ACCORDION study design was approved by Wake Forest University, USA (coordinating center) and the institutional review boards at each center (participating clinical sites). Written informed consent was obtained from all participants.

### Data collection

Upon enrollment, participants underwent questionnaires, physical examinations, and laboratory measures using a standardized protocol. Information on sociodemographic variables, lifestyle factors, and medication use were self-reported. Screened lipid levels were measured at a local laboratory or obtained from the medical records. If obtained from medical records, the most recent values recorded within the previous 12 months were used. If there were no lipid values in the medical records within the previous 12 months, the protocol specified that a blood test must be performed by the local laboratory. For participants in the trial (regardless of lipid trial assignment), post-randomization visits occurred at least every 12 months thereafter whereby serum lipids were measured using collected fasting samples.

### Calculation of cumRC

The cumRC was developed from the lipids obtained across four measurements (i.e., trial baseline in addition to the first three follow-up visits). Patients were excluded from the study if their baseline (screening) RC values were missing or if they had fewer than three valid RC measurements during follow-up. Participants with a history of CVD or those who developed MACEs before the third visit were also excluded. Ultimately, a total of 5143 participants were included in the present study (Supplementary Figure 1).

RC levels at each study visit were calculated as: RC = total cholesterol – HDL-C – LDL-C [15]. As shown in Supplementary Figure 2, cumRC was defined as the summation of the average RC for each pair of consecutive examinations multiplied by the time (in years) between these two consecutive visits: cumRC = ([RC_visit 0_ + RC_visit 1_]/2 × time_0−1_) + ([RC_visit 1_ + RC_visit 2_]/2 × time_1−2_) + ([RC_visit 2_ + RC_visit 3_]/2 × time_2−3_). RC_visit 0_, RC_visit 1_, RC_visit 2_, and RC_visit 3_ indicate the RC levels at baseline and the first, second, and third examinations, respectively, and time_0–1_, time_1–2_, and time_2–3_ indicate the participant-specific time intervals between consecutive visits in years. The time points for the measurement of blood lipids and clinical follow-up were 0, 12, 24, and 36 months.

### Outcomes

The primary outcome was the occurrence of MACEs after the third follow-up visit, including nonfatal myocardial infarction (MI), nonfatal stroke, and CVD death. Cardiovascular causes of death include fatal MI, congestive heart failure, documented arrhythmia, death after invasive cardiovascular interventions, death after noncardiovascular surgery, fatal stroke, unexpected death presumed to be due to ischemic CVD occurring <24 hours after the onset of symptoms, and death due to other vascular diseases (eg, pulmonary emboli, abdominal aortic aneurysm rupture) [14].

### Statistical analysis

Baseline participant characteristics were reported according to the quartile of cumRC. Continuous data were represented as median (IQR) with intergroup differences being evaluated using the Kruskal-Wallis nonparametric test. Categorical data were depicted as frequencies (percentages) and group disparities were determined using the χ^2^ test.

The Kaplan-Meier method was used to evaluate the incidence rate of MACEs across cumRC quartile and the group differences were evaluated using a log-rank test. Cox proportional hazard regression models were used to evaluate the relationships of cumRC with the risk of MACEs and the disease subtypes. Accordingly, multivariable-adjusted hazard ratios (HRs) and 95% confidence intervals (CIs) of MACEs across quartile of cumRC (the lowest quartile as the reference) and for 1 SD increment of cumRC were calculated. Three multivariable models were constructed with an increasing degree of covariate adjustment. Model 1 was adjusted for age (years), sex, education (less than high school, high school graduate or GED, some college, college degree or higher), smoking status (current, former, never), drinking status (0, 1-5, 6-10, >10 g/week), systolic BP (mmHg), and heart rate (bpm). Model 2 was further adjusted for estimated glomerular filtration rate (eGFR) (ml/min·1.73m^2^), HbA1c (%), and LDL-C (mg/dL). The full model (model 3) was further adjusted for assignment of the trials, including the glycemia trial (intensive, standard), the lipid trial (fenofibrate, placebo), and the BP-lowering trial (intensive, standard). P-for-trend tests were performed by treating the median cumRC values for each quartile as a continuous variable.

A multivariable restricted cubic spline regression model was used to assess the potential nonlinearity for the examined associations, with the full adjustment for the aforementioned covariates in model 3. Stratified analyses were conducted based on age (< 65 years vs. ≥ 65 years), sex (male vs. female), and assignment of the lipid trial (fenofibrate vs. placebo). To examine whether the association of cumRC with total or specific MACEs was independent of baseline RC, a supplementary analysis was performed by additionally adjusting for baseline levels of RC based on model 3.

All analyses were executed using Stata version 16 and R version 4.3.2 (R Foundation for Statistical Computing), with a 2-tailed P value <0.05 being considered statistically significant.

## Results

### Participant characteristics

Median age for the 5143 included participants was 61.7 (IQR: 58.0-66.3) years and 56.8% were male. Participants with higher levels of cumRC were more likely to be male and ethnically white, were less likely to have a college or higher degree, and were more likely to be in the intensive arm of the BP trial, the standard arm of the glycemia trial, and the placebo arm of the lipid trial. Higher cumRC levels were also associated with higher levels of diastolic BP, eGFR, total cholesterol, triglycerides, and fasting plasma glucose and lower levels of HDL-C **(Table 1)**.

**Table 1.** Baseline characteristics of participants by cumulative remnant cholesterol quartiles.

|  | Total | Quartile for cumulative remnant cholesterol |  |  |  | *P*-value |
| --- | --- | --- | --- | --- | --- | --- |
|  |  | Q1<br>(11.5-61.5) | Q2<br>(62-86.5) | Q3<br>(87-122.5) | Q4<br>(>123) |  |
| **Participants** | 5143 | 1300 | 1299 | 1281 | 1263 |  |
| **Age, years** | 61.7 (58.0-66.3) | 62.4 (58.3-67.3) | 61.9 (57.9-66.6) | 61.5 (58.0-66.2) | 61.1 (57.7-65.3) | 0.605 |
| **Male** | 2944 (56.80) | 746 (56.90) | 730 (55.90) | 698 (54.20) | 770 (60.30) | **0.015** |
| **Education** |  |  |  |  |  | **0.002** |
| Less than high school | 1440 (28.01) | 355 (27.23) | 366 (28.25) | 379 (29.53) | 340 (27.04) |  |
| High school graduate<br>or GED | 1674 (32.49) | 399 (30.66) | 421 (32.39) | 395 (30.85) | 459 (36.13) |  |
| Some college | 1344 (26.20) | 342 (26.32) | 322 (24.81) | 348 (27.27) | 332 (26.41) |  |
| College degree or<br>higher | 682 (13.24) | 202 (15.64) | 189 (14.47) | 159 (12.35) | 132 (10.42) |  |
| **Race/Ethnicity** |  |  |  |  |  | **<0.001** |
| White | 3236 (62.93) | 596 (45.84) | 777 (59.95) | 894 (69.77) | 969 (76.65) |  |
| Black | 949 (18.46) | 488 (37.53) | 250 (19.22) | 139 (10.80) | 72 (5.80) |  |
| Hispanic | 347 (6.72) | 95 (7.32) | 110 (8.42) | 82 (6.37) | 60 (4.70) |  |
| Other | 611 (11.89) | 121 (9.31) | 162 (12.40) | 166 (13.05) | 162 (12.85) |  |
| **Smoking status** |  |  |  |  |  | 0.203 |
| Never | 2478 (48.09) | 656 (50.42) | 634 (48.70) | 618 (48.25) | 570 (44.91) |  |
| Former | 2151 (41.78) | 517 (39.66) | 533 (41.12) | 538 (41.88) | 563 (44.51) |  |
| Current | 514 (10.14) | 127 (9.92) | 132 (10.18) | 125 (9.87) | 130 (10.58) |  |
| **Alcohol consumption, g/week** |  |  |  |  |  | 0.484 |
| 0 | 3850 (74.85) | 960 (73.76) | 958 (73.81) | 979 (76.30) | 953 (74.90) |  |
| 1-5 | 988 (19.21) | 255 (19.68) | 271 (20.83) | 231 (18.03) | 231 (18.10) |  |
| 6-10 | 192 (3.71) | 52 (3.97) | 47 (3.60) | 41 (3.19) | 52 (4.04) |  |
| >10 | 113 (2.24) | 33 (2.59) | 23 (1.76) | 30 (2.49) | 27 (2.10) |  |
| **SBP, mmHg** | 135.0 (125.0-146.0) | 134.0 (124.0-146.0) | 135.0 (124.0-146.0) | 135.0 (125.0-146.0) | 136.0 (125.0-148.0) | 0.500 |
| **DBP, mmHg** | 75.0 (69.0-82.0) | 74.0 (68.0-81.0) | 75.0 (69.0-82.0) | 76.0 (70.0-83.0) | 76.0 (70.0-83.0) | **0.028** |
| **Heart rate, bpm** | 73.0 (66.0-81.0) | 72.0 (65.0-80.0) | 73.0 (66.0-81.0) | 74.0 (66.0-82.0) | 75.0 (67.0-82.0) | 0.188 |
| **eGFR, ml/min·1.73m<sup>2</sup>** | 90.0 (76.9-105.2) | 90.9 (78.7-105.9) | 89.6 (76.5-104.8) | 89.6 (75.7-105.2) | 89.7 (76.1-105.0) | **0.020** |
| **TG, mg/dL** | 154.0 (108.0-227.0) | 89.0 (69.0-113.0) | 137.0 (111.0-170.0) | 183.0 (145.0-232.0) | 273.0 (206.0-367.0) | **<0.001** |
| **TC, mg/dL** | 180.0 (156.0-208.0) | 168.0 (147.0-195.0) | 177.0 (155.0-204.0) | 184.0 (160.0-211.0) | 193.0 (167.0-226.5) | **<0.001** |
| **HDL-C, mg/dL** | 41.0 (35.0-49.0) | 48.0 (40.0-56.0) | 42.0 (37.0-49.0) | 39.0 (34.0-46.0) | 36.0 (31.0-42.0) | **<0.001** |
| **LDL-C, mg/dL** | 102.0 (82.0-126.0) | 101.0 (82.0-124.0) | 104.0 (85.0-128.0) | 105.0 (83.0-128.0) | 99.0 (78.0-125.0) | 0.070 |
| **HbA1c, mmol/mol** | 64.0 (58.0-73.0) | 64.0 (58.0-73.0) | 64.0 (58.0-72.0) | 64.0 (58.0-73.0) | 65.0 (58.0-73.0) | 0.463 |
| **HbA1c, %** | 8.0 (7.5-8.8) | 8.0 (7.5-8.8) | 8.0 (7.5-8.7) | 8.0 (7.5-8.8) | 8.1 (7.5-8.8) | 0.463 |
| **FPG, mg/dL** | 167.0 (139.0-203.0) | 156.0 (131.0-193.0) | 165.0 (136.0-199.0) | 171.0 (142.0-207.0) | 176.0 (148.0-215.0) | **0.015** |
| **Glycemia Trial** |  |  |  |  |  | **0.002** |
| Intensive | 2532 (49.23) | 688 (52.92) | 648 (49.88) | 619 (48.32) | 577 (45.68) |  |
| Standard | 2611 (50.77) | 612 (47.08) | 651 (50.12) | 662 (51.68) | 686 (54.32) |  |
| **BP Trial** |  |  |  |  |  | **0.004** |
| Intensive | 1140 (22.17) | 272 (20.92) | 274 (21.09) | 267 (20.84) | 327 (25.89) |  |
| Standard | 4003 (77.83) | 1029 (79.08) | 1025 (78.91) | 1014 (79.16) | 936 (74.11) |  |
| **Lipid Trial** |  |  |  |  |  | **<0.001** |
| Fenofibrate | 1429 (27.79) | 429 (33.00) | 415 (31.95) | 340 (26.54) | 245 (19.40) |  |
| Placebo | 3714 (72.21) | 871 (67.00) | 884 (68.05) | 941 (73.46) | 1018 (80.60) |  |

Data are presented as median (P25-P75) range for continuous variables and n (%) for categorical variables.
BP, blood pressure; DBP, diastolic blood pressure; eGFR, estimated glomerular filtration rate; FPG, fasting plasma glucose; HDL-C, high-density lipoprotein cholesterol; LDL-C, low-density lipoprotein cholesterol; SBP, systolic blood pressure; TC, total cholesterol; TG, triglycerides.

### Associations of cumRC levels with risk of MACEs

During a median follow-up period of 6.3 years, 472 participants developed MACEs, including 154 CVD deaths, 211 with nonfatal MI, and 148 with nonfatal stroke. Participants in the higher quartiles of cumRC tended to have a higher cumulative incidence of MACEs, according to the log-rank tests (P < 0.01; **Figure 1**).

**Figure 1.**
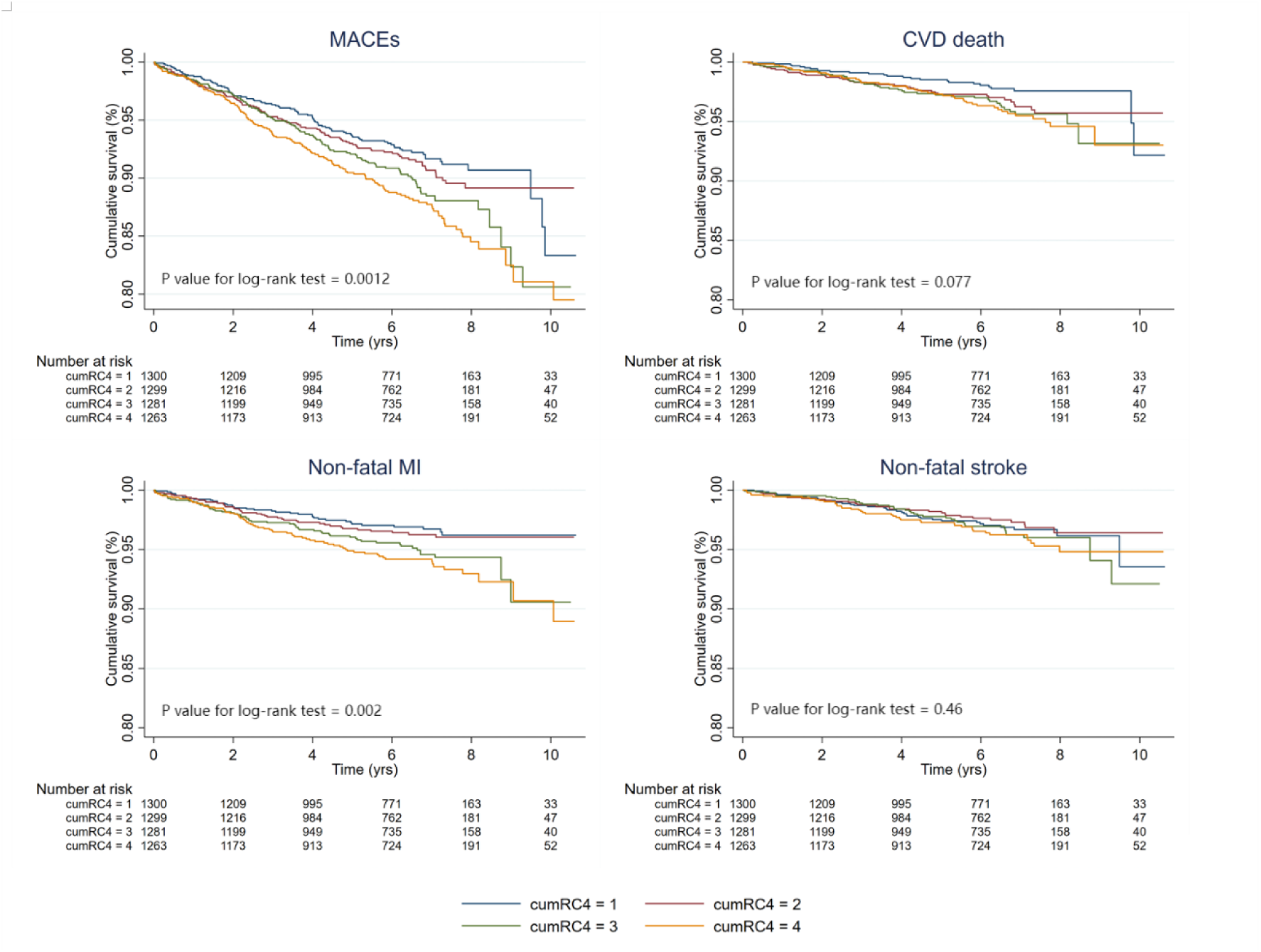
Kaplan–Meier survival curves for MACEs and individual outcomes based on quartile of cumulative remnant cholesterol.

After the multivariable adjustment, higher levels of cumRC were associated with a higher risk of MACEs (HR_Q4 vs. Q1_ = 1.71, 95% CI: 1.31-2.22; P-trend <0.001), with similar associations for CVD deaths and nonfatal MI, but no association for nonfatal stroke (**Table 2**). The fully-adjusted HRs comparing the highest quartile of cumRC with the lowest quartile were 2.02 (95% CI: 1.23-3.32) for CVD death (P-trend = 0.009), 2.03 (95% CI: 1.36-3.04) for nonfatal MI (P-trend <0.001), and 1.34 (95% CI: 0.86-2.11) for nonfatal stroke (P-trend = 0.099).

**Table 2.**
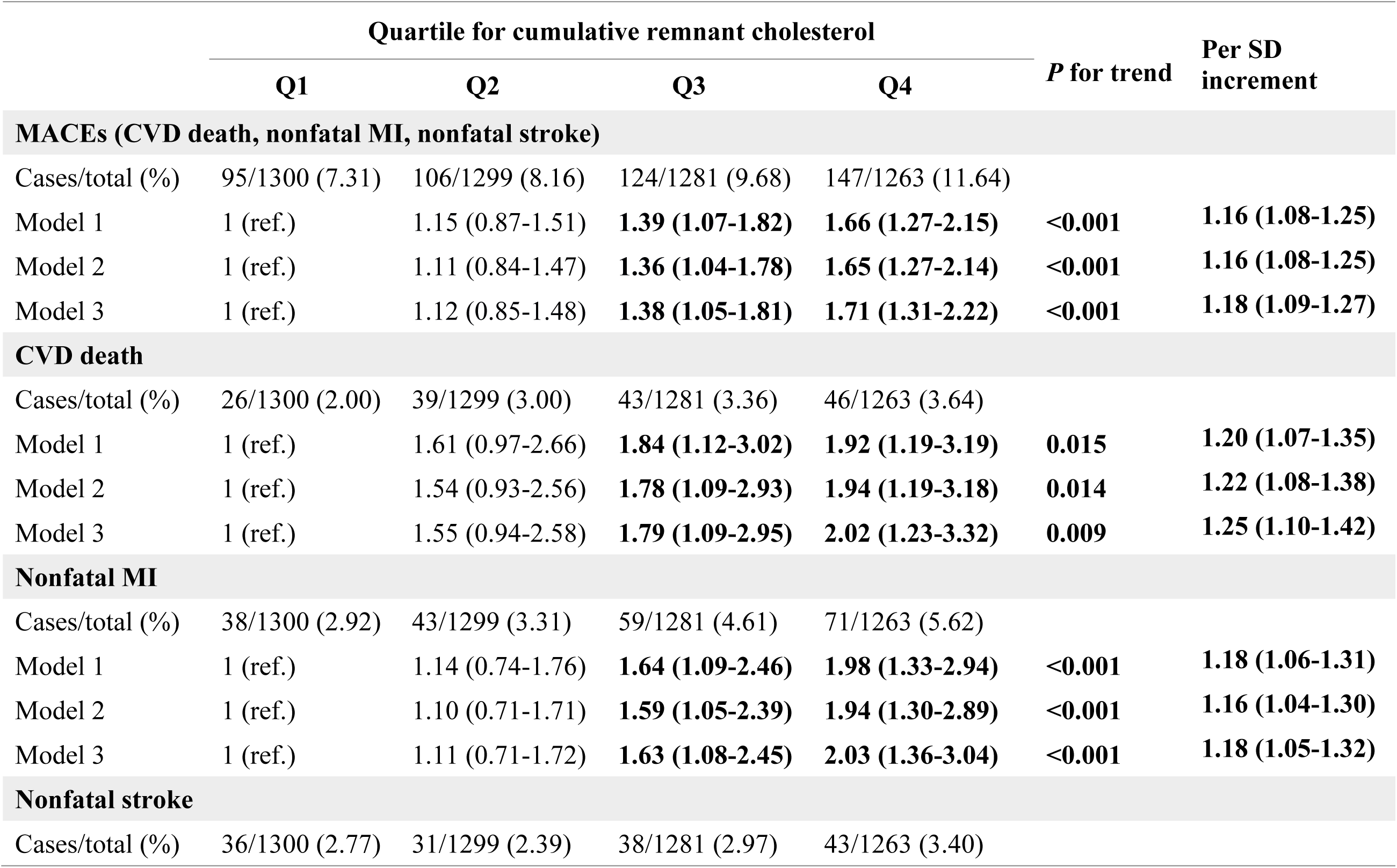
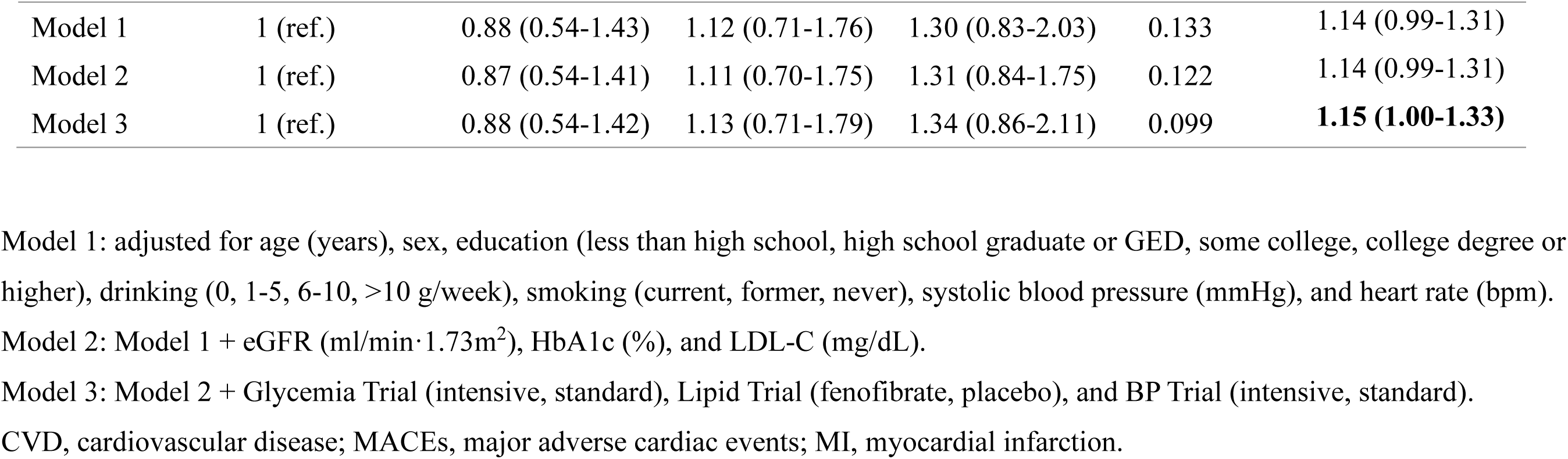
The associations of cumulative remnant cholesterol with MACEs and individual cardiovascular diseases.

Including cumRC as a continuous variable in the full model, each 1-SD increase of cumRC was associated with an 18% (95% CI: 9%-27%) higher risk of MACEs, a 25% (95% CI: 10%-42%) higher risk of CVD death, and an 18% (95% CI: 5%-32%) higher risk of nonfatal MI. There was no evidence for a nonlinear relationship between cumRC levels and the risk of total or specific MACEs (P-nonlinearity >0.20) (**Figure 2**).

**Figure 2.**
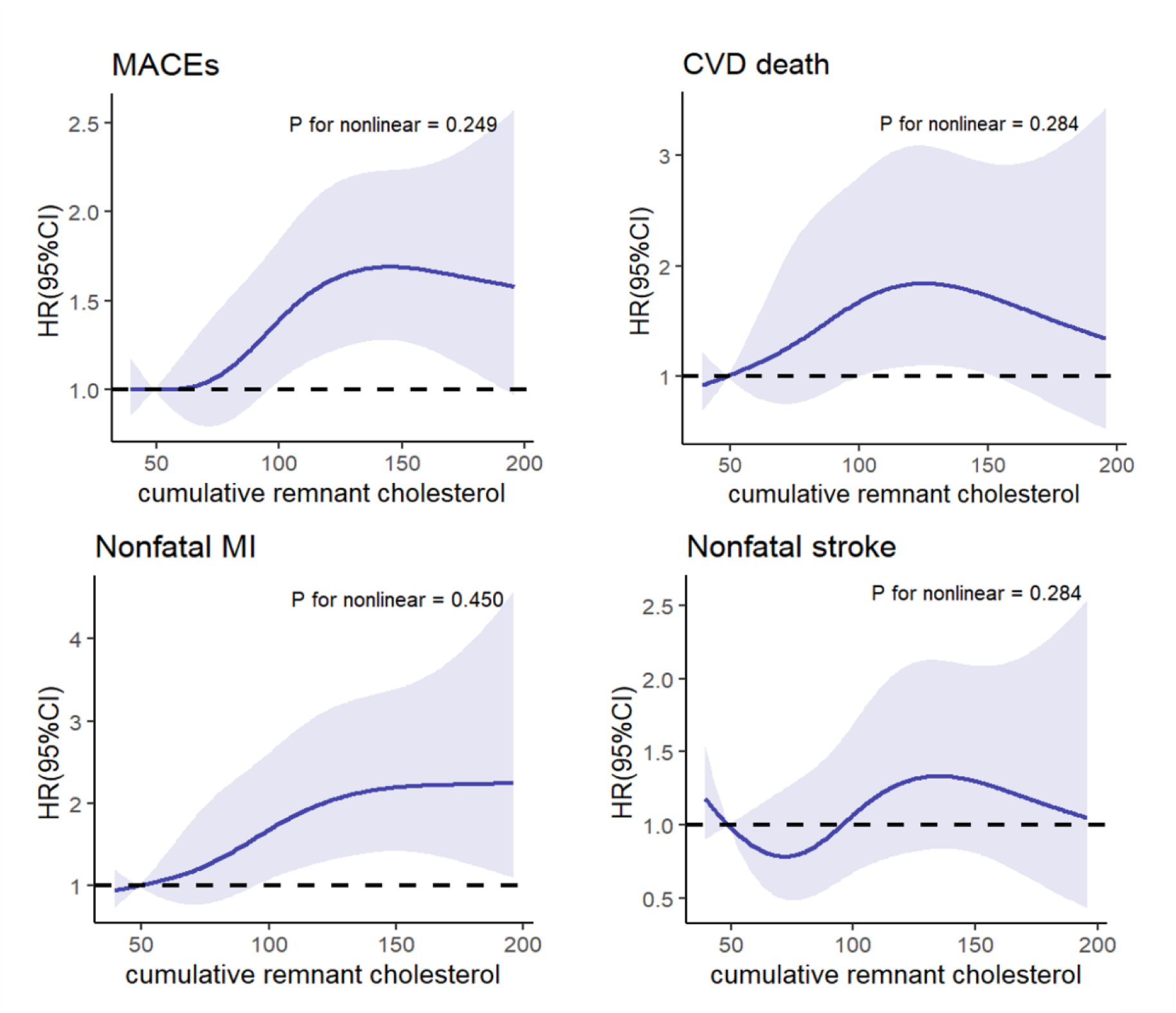
**Restricted cubic splines for the relationship of cumulative remnant cholesterol levels with risk of MACEs and individual cardiovascular diseases.** The splines were modelled with four knots (25th, 50th, 75th, and 95th percentiles), and the 12.5th percentiles was used as the reference. Results were adjusted for age (years), sex, education (less than high school, high school graduate or GED, some college, college degree or higher), drinking (0, 1-5, 6-10, >10 g/week), smoking (current, former, never), systolic blood pressure (mmHg), heart rate (bpm), eGFR (ml/min·1.73m^2^), HbA1c (%), LDL-C (mg/dL), Glycemia Trial (intensive, standard), Lipid Trial (fenofibrate, placebo), and BP Trial (intensive, standard). MACEs, major adverse cardiac events.

### Subgroup and supplementary analyses

The association of cumRC with the risk of MACEs was consistent across the subgroups defined by age (< 65 years vs. ≥ 65 years), sex (male vs. female), or lipid trial assignment (fenofibrate vs. placebo) (P-interaction >0.05) (**Figure 3**). The associations of cumRC with the risk of MACEs or specific cardiovascular outcomes were attenuated only modestly after further adjusting for baseline levels of RC (Supplementary Table 1).

**Figure 3.**
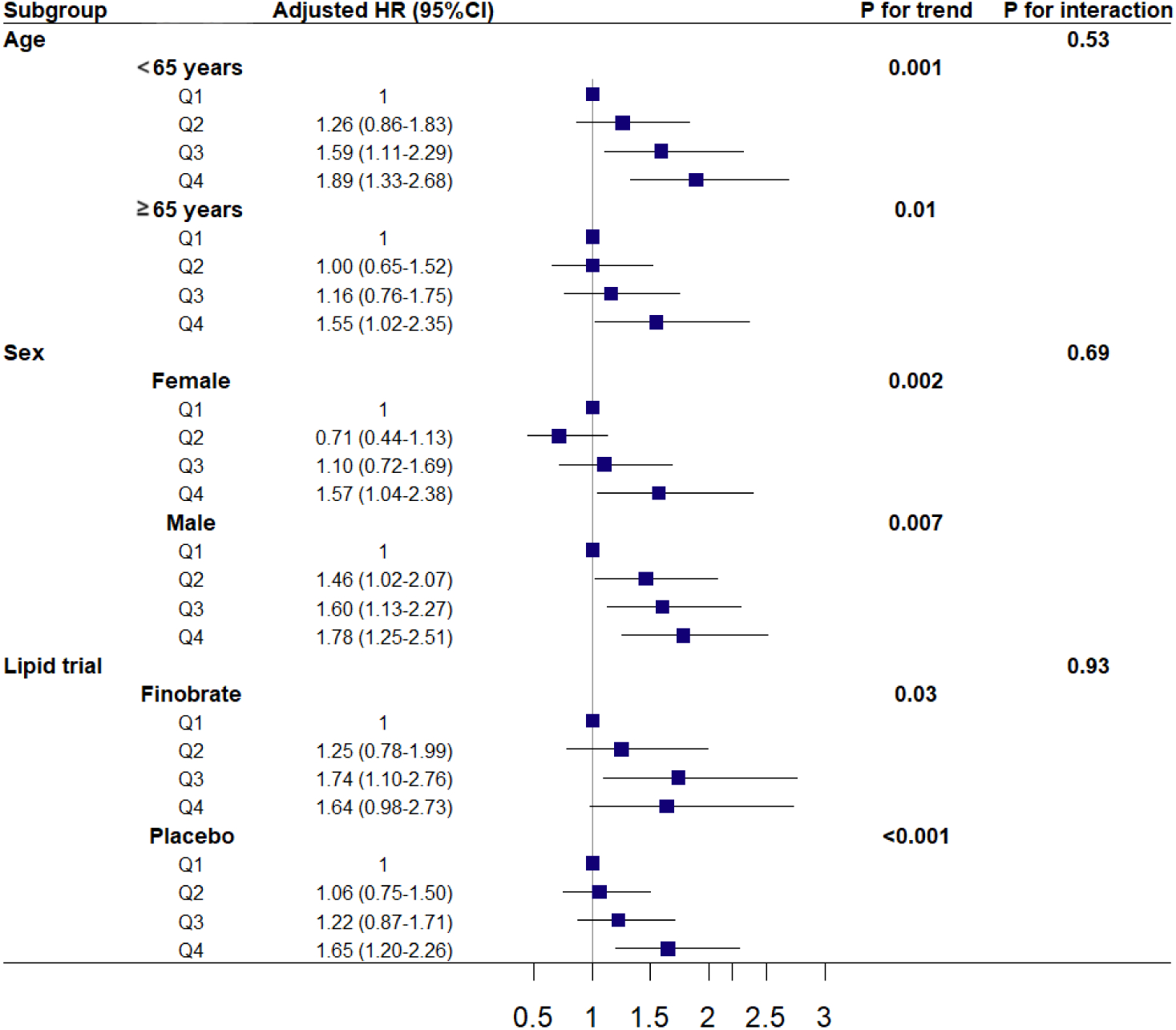
**Stratified analysis on the relationship between cumulative remnant cholesterol levels and risk of MACEs** Results were adjusted for age (years), sex, education (less than high school, high school graduate or GED, some college, college degree or higher), drinking (0, 1-5, 6-10, >10 g/week), smoking (current, former, never), systolic blood pressure (mmHg), heart rate (bpm), eGFR (ml/min·1.73m^2^), HbA1c (%), LDL-C (mg/dL), Glycemia Trial (intensive, standard), Lipid Trial (fenofibrate, placebo), and BP Trial (intensive, standard). BP, blood pressure; eGFR, estimated glomerular fltration rate; HbA1c, glycated hemoglobin; LDL-C, low-density lipoprotein cholesterol, MACEs, major adverse cardiovascular events.

## Discussion

Individuals with type 2 diabetes are more susceptible to dyslipidemia and atherosclerotic CVD than the general population. In this post hoc analysis of the ACCORD study, we examined the relationship between 3-year cumulative levels of RC and the subsequent risk of developing MACEs among adults with type 2 diabetes. We found that higher levels of cumRC were associated with a higher risk of developing MACEs, after adjusting for traditional cardiovascular risk factors. This association was similar across age, sex, and lipid trial assignment groups and was independent of RC levels assessed at baseline. Higher cumRC levels were also associated with a higher risk of cardiovascular death and nonfatal MI, but not nonfatal stroke. These findings emphasize the importance of dynamic monitoring and management of RC among individuals with type 2 diabetes, even in the context of intensive lipid-lowering treatment.

Many previous studies, largely conducted in the general population with a single lipid measurement, have shown that elevated RC levels were associated with an increased risk of CVD, independently of LDL-C levels [16–18]. These findings suggested that the atherogenic effect of RC may explain the relatively higher incidence of MACEs among hyperlipidemic individuals whose LDL-C levels have been pharmacologically treated to an optimal range. The residual cardiovascular risk is particularly common for individuals with type 2 diabetes [19]. In a previous study including type 2 diabetes participants with a single assessment of RC, higher RC levels were associated with a higher risk of cardiovascular mortality and incident diabetic nephropathy [20]. In the present study, the relationship between cumRC and elevated risk of MACEs remained among participants who received intensive lipid-lowering therapies. This further supports that cumulative exposure to elevated RC may be an important contributor to the widely observed residual risk for atherosclerotic CVD on the basis of pharmacological lowering of LDL-C levels.

The majority of the previous studies assessing the RC-CVD association have been focused on a single measurement of RC levels while ignoring the effect of cumulative exposure. Cardiovascular damage evolves over prolonged periods, and so a single assessment of risk factors is suboptimal as a measurement proxy for long-term exposure. Furthermore, within person variability of the measurement may further bias the relationship between baseline risk factors and future CVD risk. Some studies have shown that, as compared to the exposure to certain cardiovascular risk factors (e.g., higher BP) assessed at a single time point, long-term cumulative exposure has a more pronounced adverse cardiovascular effect [21]. Wu et al. [22] found that increased cumRC exposure was associated with a higher risk of CVD among Chinese participants with hypertension, even after adjusting for baseline RC measurements. A special report by the American Heart Association and the American College of Cardiology states that the majority of heart attack and stroke events are attributable to cumulative exposure to preventable disease-causing risk factors [23]. Taken together, our findings suggest that the cardiovascular impact of RC among individuals with type 2 diabetes is jointly determined by the magnitude and the cumulative duration of exposure.

The pathophysiologic process of atherosclerosis may explain the observed association between cumRC and MACEs. Similar to LDL cholesterol, RC has been recognized as a potential contributor to atherosclerosis [24, 25]. Circulating lipoproteins that cross the endothelial barrier can interact with extracellular structures and are retained in the extracellular matrix [26, 27]. High serum concentrations of RC have the ability to penetrate the arterial wall, tend to accumulate within the vessel wall, and appear to be more readily taken up by smooth muscle cells and macrophages than LDL-C [28, 29], thereby inducing maladaptive and complex inflammatory processes that lead to the subsequent development of atherosclerosis [30].

Our study may have important clinical implications. In the well-defined ACCORD study, we established a longitudinal correlation between cumRC and the future risk of developing MACEs in a type 2 diabetes population, with a similar association among those who were intensively treated for LDL-C lowering. These findings may provide new insights into the pathogenesis of residual cardiovascular risk and refine the methods for CVD risk assessment in this high-risk population group. By incorporating the cumRC burden into clinical research of type 2 diabetes, our findings may further guide the prevention and therapy strategies for MACEs among adults with type 2 diabetes. Future research is needed to assess whether the adoption of behavioral (e.g., healthy diet and lifestyle) or pharmacological therapies that target RC levels can further slow the progression of atherosclerosis and prevent cardiovascular events among adults with type 2 diabetes.

There are a few potential limitations to this study. First, the majority of the participants were ethnically white, which may limit the generalization of our findings to other racial/ethnic populations. Second, despite adjusting for other known cardiovascular risk factors, the potential influence of residual confounding on our results cannot be completely ruled out. Third, our study is a post hoc rather than prespecific analysis of the ACCORD trial. A significant number of participants were excluded from the analysis (e.g., with fewer than three measurements of RC levels in the first three years), which may lead to potential selection bias. Finally, the population of the ACCORD trial included only high-risk type 2 diabetes patients, a part of whom were undergoing intensive glycemic or lipid therapies. Although we have controlled for these variables, more studies are needed to improve the generalizability of the results.

## Conclusions

In a post hoc analysis of the ACCORD trial of type 2 diabetes participants, our findings suggest that cumulative exposure to elevated RC is associated with a higher risk of developing MACEs, independently of known risk factors and baseline RC levels. In particular, the association remained among type 2 diabetes participants who were undergoing intensive lipid-lowering treatment. These findings emphasize the importance of not only regular monitoring of RC, but also enhanced assessment of cumulative RC levels, to identify those with residual risk for MACEs in spite of successful LDL-C management.

## Data Availability

All data produced are available online at https://biolincc.nhlbi.nih.gov/studies/accord/

## Acknowledgments

The authors gratefully acknowledge the ACCORD study group and the National Heart, Lung, and Blood Institute Biologic Specimen and Data Repository Information Coordinating Center, for conducting the trials and making data sets publicly available. The opinions expressed in this article are those of the authors and do not necessarily reflect the views of the ACCORD study authors or the National Heart, Lung, and Blood Institute.

## Availability of data and materials

The datasets used and analyzed during the current study are available from the ACCORD/ACCORDION Research Materials obtained from the National Heart, Lung, and Blood Institute (NHLBI) Biologic Specimen and Data Repository Information Coordinating Center. The contents of this report do not necessarily reflect the opinions or views of the ACCORD/ACCORDION study authors or the NHLBI.

## Abbreviations

ACCORD: the Action to Control Cardiovascular Risk in Diabetes
CVD: cardiovascular disease
DBP: diastolic blood pressure
eGFR: estimated glomerular filtration rate
FPG: fasting plasma glucose
HDL-C: high-density lipoprotein cholesterol
LDL-C: low-density lipoprotein cholesterol
MACEs: major adverse cardiovascular events
MI: myocardial infarction
RC: remnant cholesterol
SBP: systolic blood pressure
TC: total cholesterol
TG: triglycerides

